# COVID-19 due to the B.1.617.2 (Delta) variant compared to B.1.1.7 (Alpha) variant of SARS-CoV-2: two prospective observational cohort studies

**DOI:** 10.1101/2021.11.24.21266748

**Authors:** Kerstin Kläser, Erika Molteni, Mark Graham, Liane Canas, Marc FÖsterdahl, Michela Antonelli, Liyuan Chen, Jie Deng, Benjamin Murray, Eric Kerfoot, Jonathan Wolf, Anna May, Ben Fox, Joan Capdevila, The COVID-19 Genomics UK (COG-UK) consortium, Marc Modat, Alexander Hammers, Tim D. Spector, Claire J. Steves, Carole H. Sudre, Sebastien Ourselin, Emma L. Duncan

**Author notes:** **Corresponding author** Emma L Duncan, Professor of Clinical Endocrinology, Department of Twin Research & Genetic Epidemiology, School of Life Course Sciences, Faculty of Life Sciences and Medicine, King’s College London, Mailing Address: Department of Twin Research, King’s College London, St Thomas’ Hospital Campus, 3rd Floor South Wing Block D, Westminster Bridge Road, London SE1 7EH. Contributed equally (Sebastien Ourselin, Emma L. Duncan). **Ethics** Ethics approval was granted by KCL Ethics Committee (reference LRS-19/20-18210).

## Abstract

**Background:** The Delta (B.1.617.2) variant became the predominant UK circulating SARS-CoV-2 strain in May 2021. How Delta infection compares with previous variants is unknown.

**Methods:** This prospective observational cohort study assessed symptomatic adults participating in the app-based COVID Symptom Study who tested positive for SARS-CoV-2 from May 26 to July 1, 2021 (Delta overwhelmingly predominant circulating UK variant), compared (1:1, age- and sex-matched) with individuals presenting from December 28, 2020 to May 6, 2021 (Alpha (B.1.1.7) predominant variant). We assessed illness (symptoms, duration, presentation to hospital) during Alpha- and Delta-predominant timeframes; and transmission, reinfection, and vaccine effectiveness during the Delta-predominant period.

**Findings:** 3,581 individuals (aged 18 to 100 years) from each timeframe were assessed. The seven most frequent symptoms were common to both variants. Within the first 28 days of illness, some symptoms were more common with Delta vs. Alpha infection (including fever, sore throat and headache) and vice versa (dyspnoea). Symptom burden in the first week was higher with Delta vs. Alpha infection; however, the odds of any given symptom lasting ≥7 days was either lower or unchanged. Illness duration ≥28 days was lower with Delta vs. Alpha infection, though unchanged in unvaccinated individuals. Hospitalisation for COVID-19 was unchanged. The Delta variant appeared more (1·47) transmissible than Alpha. Re-infections were low in all UK regions. Vaccination markedly (69-84%) reduced risk of Delta infection.

**Interpretation:** COVID-19 from Delta or Alpha infections is clinically similar. The Delta variant is more transmissible than Alpha; however, current vaccines show good efficacy against disease.

**Funding:** UK Government Department of Health and Social Care, Wellcome Trust, UK Engineering and Physical Sciences Research Council, UK Research and Innovation London Medical Imaging & Artificial Intelligence Centre for Value Based Healthcare, UK National Institute for Health Research, UK Medical Research Council, British Heart Foundation, Alzheimer’s Society, and ZOE Limited.

**Research in context:** *Evidence before this study:* To identify existing evidence for differences (including illness, transmissibility, and vaccine effectiveness) from SARS-CoV-2 infection due to Alpha (B.1.1.7) and Delta (B.1.617.2) variants, we searched PubMed for peer-reviewed articles and medRxiv for preprint publications between March 1 and November 18, 2021 using keywords (“SARS-CoV-2” OR “COVID-19”) AND (“delta variant” OR “B.1.617.2”) AND (symptom* OR transmiss* OR “disease duration” OR “illness duration” OR “symptom* duration”). Searches were not restricted by language. Among 169 identified PubMed articles, we found evidence that Delta variant has increased replication capacity (from 4-fold, up to 21-fold, compared with wild-type) and greater transmissibility (estimated between +20% and +97%), compared with previous strains. Currently available vaccines may have 2- to 5-fold lower neutralizing response to Delta vs. previous variants, depending on vaccine formulation, although their protective effect against severe disease and death appears to remain strong. REACT-1 study found that in UK infections were increasing exponentially in the 5-17-year old children in September 2021, coinciding with the start of the autumn school term in England. This was interpreted as an effect of the relatively low rate of vaccinated individuals in this age group. Other studies found that in unvaccinated individuals, Delta variant may be associated with higher odds of pneumonia, oxygen requirement, emergency care requests, ICU admission, and death. In a study of 27 (mainly young) cases, 22 persons were symptomatic, with fever (41%), cough (33%), headache (26%), and sore throat (26%) the commonest symptoms. We found no studies, beyond case series, investigating symptom and/or illness duration due to Delta variant infection otherwise.

*Added value of this study:* Using data from one of the largest UK citizen science epidemiological initiatives, we describe and compare illness (symptom duration, burden, profile, risk of long illness, and hospital attendance) in symptomatic community-based adults presenting when either the Alpha or Delta variant was the predominant circulating strain of SARS-CoV-2 in the UK. We assess evidence of transmission, reinfection, and vaccine effectiveness. Our data show that the seven most common symptoms with Delta infection were the same as with Alpha infection. Risks of illness duration ≥7 days and ≥28 days, and of requiring hospital care, were not increased. In line with previous research, we found increased transmissibility of Delta vs. previous variants; and no evidence of increased re-infection rates. Our data support high vaccine efficacy of BNT162b2 and ChAdOx1 nCoV-19 formulations against Delta variant infection. Overall, our study adds quantitative information regarding meaningful clinical differences in COVID-19 due to Delta vs. other variants.

*Implications of all the available evidence:* Our observational data confirm that COVID-19 disease in UK in adults is generally comparable to infection with the Alpha variant, including in elderly individuals. Our data contribute to epidemiological surveillance from the wider UK population and may capture information from COVID-19 presentation within the community that might be missed in healthcare-based surveillance. Our data may be useful in informing healthcare service planning, vaccination policies, and measures for social protection.

## Introduction

Viruses mutate over time,^1^ affecting transmissibility,^2^ disease presentation,^3^ and natural or vaccine-induced protective immunity.^4^ The Delta variant of SARS-CoV-2 (B.1.617.2) was identified in India in late 2020 and declared a ‘Variant of Concern’ in May 2021 by the UK,^5^ the World Health Organization,^6^ and the European Centre for Disease Control,^7^ mainly due to evidence of increased transmissibility,^8,9^ possibly larger risk of hospitalisation^10,11^ and conceivably less effectiveness of vaccination compared with previous variants.^4,12,13^

In the UK, the Delta variant rapidly became the dominant circulating form of SARS-CoV-2, (from 0·09% at the beginning of April 2021 to >98% at the end of June 2021) displacing the Alpha (B.1.1.7) variant which concomitantly decreased from 98% to 1·67% (Supplementary Table S1, Supplementary Figure S1).^14^ Many other factors also changed contemporaneously, including SARS-CoV-2 prevalence, delivery of an age-stratified mass vaccination campaign, relaxation of lockdown, and test access criteria.

We previously described COVID-19 profile, transmissibility, re-infection risk,^15^ and vaccine effectiveness^16^ when the Alpha (B.1.1.7) variant was predominant. Here, we present these data after the Delta variant became predominant, and compare illness from Delta vs. Alpha infection, in a large UK community cohort.

## Methods

### COVID Symptom study

Prospective longitudinal observational data were collected as part of the King’s College London [KCL]/ZOE COVID Symptom Study, using the ZOE COVID Study App^17^ (Supplementary Information). Briefly, upon enrolment users provide baseline demographic and health information, and subsequently are prompted daily to record symptoms (or their absence) through direct questioning (Supplementary Table S2) and free text, any SARS-CoV-2 testing and corresponding result, vaccination details, and any hospital presentation. Users can also proxy-report for others. The current study was drawn from approximately 1 million UK app users who logged data at least once between December 28, 2020 to July 1, 2021.

Ethics approval was granted by the KCL Ethics Committee (LRS-19/20-18210). At registration, all participants provided consent for their data to be used for COVID-19 research. Governance was specifically granted for use of proxy-reported data.

Data from all UK adult participants aged 18 to 100 years (including proxy-reported individuals) who logged a positive PCR or lateral flow antigen test (LFAT) for SARS-CoV-2 between December 28, 2020 to July 1, 2021 were considered. As previously,^18^ individuals were considered to have COVID-19 if SARS-CoV-2-associated symptoms were reported (or proxy-reported) (Supplementary Table S2) between two weeks before and one week after positive testing. Data were included for individuals who reported at least weekly, from first symptom report until returning to symptom-freedom or until reporting ceased.^19^

Data were compared between two time periods: December 28, 2020 to May 6, 2021, when the Alpha variant was the predominant circulating SARS-CoV-2 strain (proportion of sequenced strains: >75% from December 28, 2020, reaching >95% by February 3 2021, and remaining >75% until 28 April); and May 26, 2021 to July 1, 2021, when the Delta variant was the predominant strain (>75% from May 26, reaching >95% by June 9 and >99% from 30 June to date) (Supplementary Table S1). Individuals logging a positive test did not have variant confirmation by sequencing; thus, illness within these two timeframes was attributed to the predminant circulating variant. Terminology herein reflects this assumption.

Through an Euclidean distance-based algorithm,^20^ individuals with Delta infection were matched 1:1, based on their age and sex, with individuals with Alpha infection. We were unable to match for SARS-CoV-2 prevalence, tiered lockdown restrictions, or vaccination rates, which varied widely across the community and with time during this study.

Symptom data were censored at August 5, 2021, 35 days after last inclusion date for testing positive with Delta infection, allowing at least 28 days’ symptom evaluationfor all individuals. Symptoms were considered over the entire illness, which by virtue of illness definition could extend outside SARS-CoV-2 testing date boundaries (a maximum of two weeks before and five weeks after testing, allowing for individuals whose illness started up to a week after positive test). To allow for symptom waxing and waning, individuals who returned a healthy report but subsequently logged as symptomatic within seven days of their last unhealthy report were considered unwell from their initial illness, with per-symptom and illness duration calculated accordingly. We ascertained odds of a given symptom developing within 28 days of illness; and odds of each symptom lasting ≥7 days, corrected for age, sex and vaccination status (unvaccinated, 1 dose, 2 doses), with a given vaccination considered valid after 14 days (allowing for evolving immunity). We used false discovery rate to account for multiple comparisons. We assessed risk of illness duration ≥28 days (LC28) and hospital presentation (admission or emergency room attendance), overall and for unvaccinated individuals, similarly adjusted for age, sex and vaccination status.

### Transmissibility

We used data from COVID-19 Genomics UK Consortium (COG-UK) to extract time-series of the percentage of daily positive SARS-CoV-2 testing from the Delta lineage in Scotland, Wales, and each of nine National Health Service (NHS) regions in England. Northern Ireland was excluded due to low sample numbers in the COG-UK dataset. The COG-UK data are produced by sequencing a random sample of positive PCR tests from the general community.

Daily SARS-CoV-2 incidence data for Scotland, Wales, and each NHS region in England were estimated fromMarch 14 to August 8, 2021, using CSS app data and previously described methodology.^21^ The method uses both positive SARS-CoV-2 test results and symptom reports from app users, to estimate incidence. Data are stratified by age and vaccination status to ensure estimates made from the CSS app population are representative of the wider population.

Using COG-UK data to estimate the proportion of Delta in circulation in each region per day, incidence estimates were decomposed into two incidence time-series per region, one for ‘non-Delta’ (in the timeframe considered here, predominantly Alpha [Supplementary Table S1]) and one for Delta, assuming that the two incidence time-series should sum to match total incidence. R(t) was estimated separately for non-Delta and Delta variants, using previously described methodology.^21^ Briefly, we used the relationship I_t+1_=I_t_ exp(μ (R(t) – 1)), where 1/μ is the serial interval and I_t_ the incidence on day t. We modelled the system as a Poisson process and assumed the serial interval was drawn from a gamma distribution with α=6·0 and β=1·5; and used Markov Chain Monte-Carlo to estimate R(t). We compared both multiplicative and additive differences of the new and old R(t) values for days when the Delta proportion in a region was >3%.

### Reinfection during rise of Delta variant

Reinfection was defined as previously^22^ (presence of two positive PCR or LFAT tests separated by >90 days, with an asymptomatic period of ≥7 days before the second positive test). To assess risk of reinfection during the Delta variant timeframe we performed ecological studies for each region, examining the Spearman correlation between the proportion of circulating SARS-CoV-2 due to Delta (Supplementary Table S5, Supplementary Figure S1) and number of reinfections per week over time, assessed from 10 weeks prior to Delta prevalence of 25% until 10 weeks after Delta prevalence of 75% (22 weeks); and between the number of positive tests reported through the app and the number of reinfections. We compared the bootstrapped distributions of these two correlations in each region, using the Mann-Whitney U test (Supplementary Table S5).

### Post-vaccination infection during Delta period

We analysed 515,138 app users who reported vaccination with BNT162b2 (BioNTech-Pfizer) or ChAdOx1 nCoV-19 (Oxford-Astra Zeneca) and were subsequently tested for SARS-CoV-2 (PCR or LFAT) 14-60 days after either first or second vaccination (assessed separately) after 26 May 2021.^22^ Age was restricted to 20-65 years, as most individuals >65 years were vaccinated and most individuals <20 years unvaccinated during the time of analysis, biasing the control groups for these ages. Users who had reported SARS-CoV-2 infection previously were excluded. Unvaccinated users reporting SARS-CoV-2 test results in the same or following week as a vaccinated app user served as controls. In the event of multiple tests logged for an individual vaccinated user, either the first positive or the last negative result was selected. For each vaccine and per dose, we modelled rates of positive testing in vaccinated vs. unvaccinated individuals, using Poisson regressions adjusting for number of tests, age, co-morbidities, sex, healthcare worker status, obesity, and weekly incidence in the community (by controlling for the date of the test). The adjusted risk reduction was then calculated as *RR=riskratio*_*i,n*_*-1*, where *i* is the vaccine type, and *riskratio* is the ratio of infection rates in vaccinated individuals compared to unvaccinated individuals, derived from our Poisson model.

## Results

### Cohort description

44,718 adults testing positive for SARS-CoV-2 between December 28, 2020 and July 1, 2021, 22,699 had symptoms within requisite timeframes and logged sufficiently regularly for calculation of illness duration: 19,118 individuals when Alpha was predominant and 3,581 when Delta was predominant. Demographic characteristics (after age, gender, and 1:1 matching) are presented in Table 1.

**Table 1.**
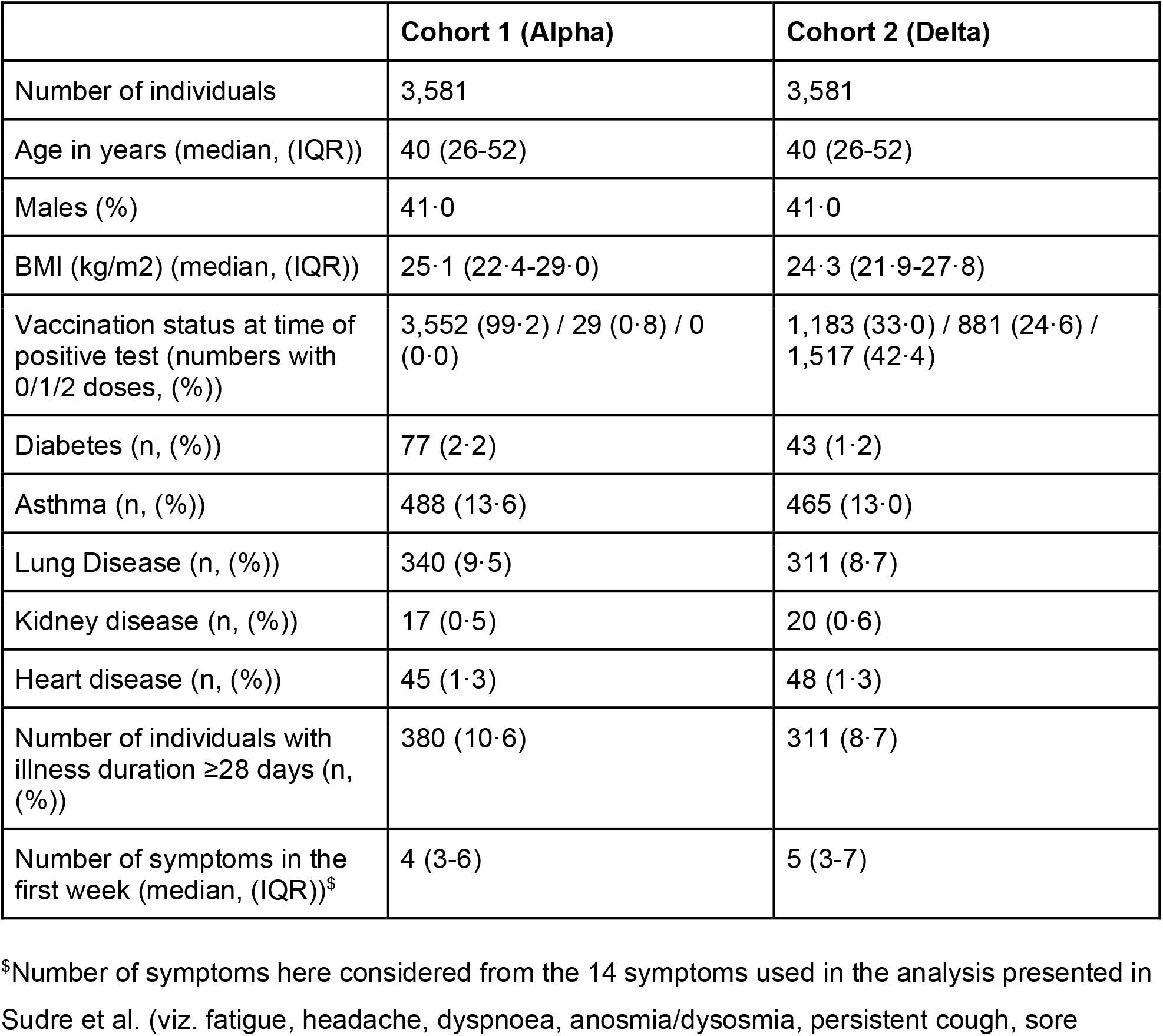
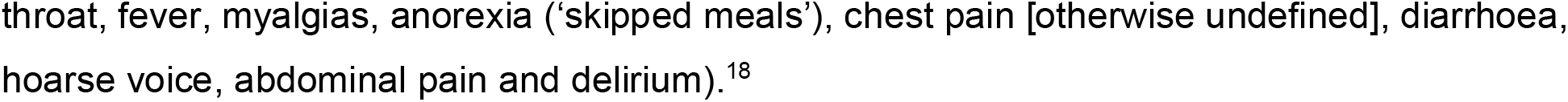
Characteristics of UK individuals presenting with COVID-19, during periods of Alpha and Delta SARS-CoV-2 variant predominance.

|  | Cohort 1 (Alpha) | Cohort 2 (Delta) |
| --- | --- | --- |
| Number of individuals | 3,581 | 3,581 |
| Age in years (median, (IQR)) | 40 (26-52) | 40 (26-52) |
| Males (%) | 41·0 | 41·0 |
| BMI (kg/m<sup>2</sup>) (median, (IQR)) | 25·1 (22·4-29·0) | 24·3 (21·9-27·8) |
| Vaccination status at time of positive test (numbers with 0/1/2 doses, (%)) | 3,552 (99·2) / 29 (0·8) / 0 (0·0) | 1,183 (33·0) / 881 (24·6) / 1,517 (42·4) |
| Diabetes (n, (%)) | 77 (2·2) | 43 (1·2) |
| Asthma (n, (%)) | 488 (13·6) | 465 (13·0) |
| Lung Disease (n, (%)) | 340 (9·5) | 311 (8·7) |
| Kidney disease (n, (%)) | 17 (0·5) | 20 (0·6) |
| Heart disease (n, (%)) | 45 (1·3) | 48 (1·3) |
| Number of individuals with illness duration $\geq 28$ days (n, (%)) | 380 (10·6) | 311 (8·7) |
| Number of symptoms in the first week (median, (IQR))<sup>\$</sup> | 4 (3-6) | 5 (3-7) |

<sup>\$</sup>Number of symptoms here considered from the 14 symptoms used in the analysis presented in Sudre et al. (viz. fatigue, headache, dyspnoea, anosmia/dysosmia, persistent cough, sore
throat, fever, myalgias, anorexia ('skipped meals'), chest pain [otherwise undefined], diarrhoea, hoarse voice, abdominal pain and delirium).<sup>18</sup>

### Illness profile

Symptoms within the first 28 days of illness are presented in Figure 1 and Supplementary Table S3 (descriptive data). All symptoms evidenced until day 28 had presented by day 21, in all individuals with both variants. The seven most reported symptoms were the same for Delta as for Alpha infection, though varied in prevalence: headache (75% vs. 67%), fatigue (73% in both), rhinorrhea (71% vs. 54%), anosmia/dysosmia (64% vs. 54%), sneezing (59% vs. 44%), sore throat (56% vs. 42%) and persistent cough (51% vs. 41%).

**Figure 1.**
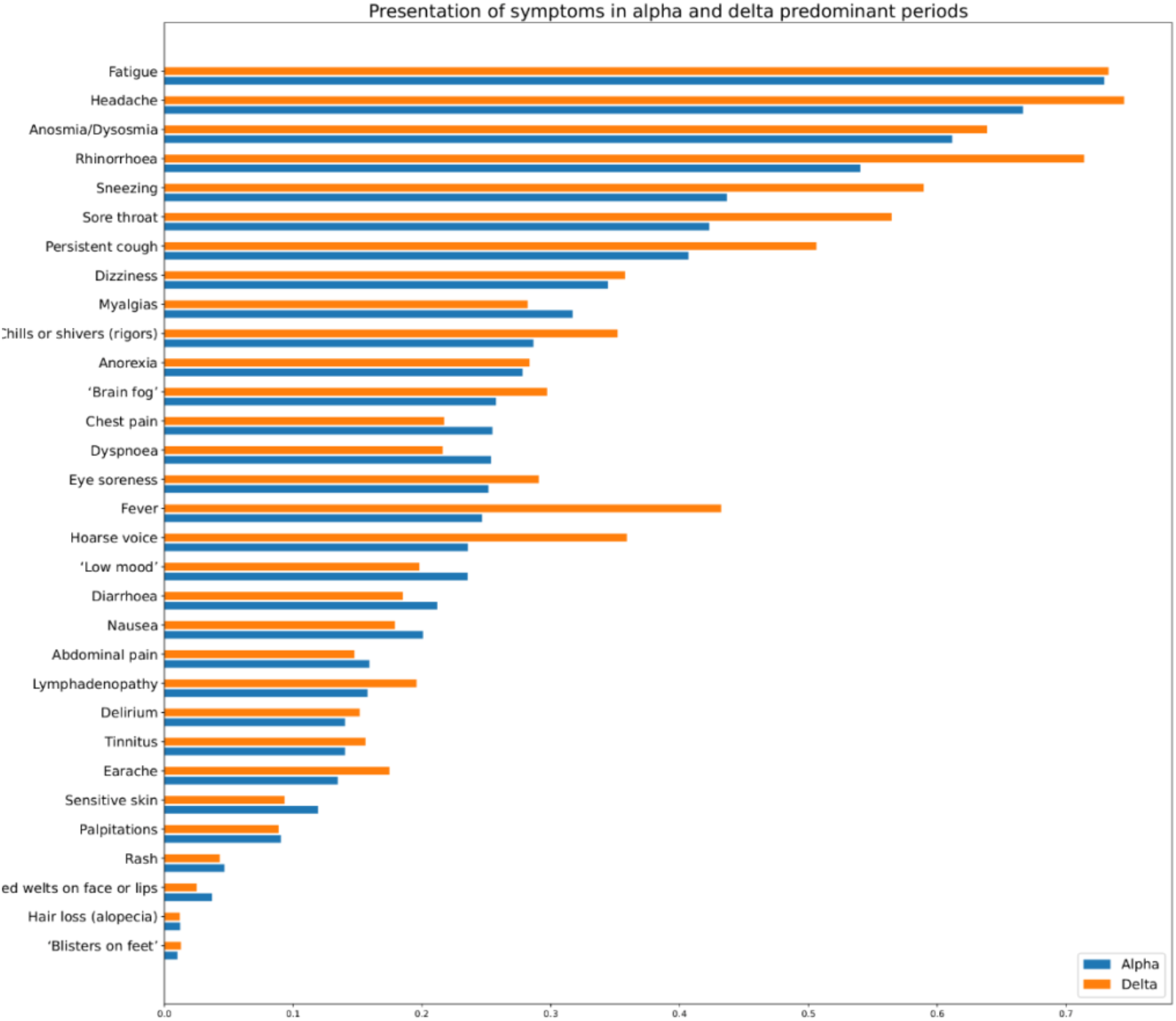
Prevalence of symptoms reported over the course of illness (up to 28 days) in individuals with COVID-19 during periods of SARS-CoV-2 Alpha or Delta variant predominance.

Correcting for age, sex, and vaccination status at time of positive test, and for false discovery rate, several symptoms were more common during the first 28 days of illness with Delta vs. Alpha infection, including fever (OR = 2·82 [95% CI 2·44-3·26]), hoarse voice (OR =1·82 [95% CI 1·56 ; 2·11]), sore throat (OR = 1·73, [95% CI 1·5 ; 2]), and persistent cough (OR = 1·64 [95% CI 1·43 ; 1·88]); conversely, the risk of shortness of breath was lower (OR = 0·82 [95% CI 0·69 ; 0·96]) (Figure 2, Supplementary Table S4). The odds of five or more symptoms in the first week of illness were higher with Delta vs. Alpha infection, whether for the 14 symptoms analysed in Sudre et al.^18^ (OR = 1·70 [95% CI: 1·47; 1·95], p<0·00005) or all symptoms (Supplementary Table S2) (OR = 1·78 [95% CI: 1·50; 2·11], p<0·00005). However, the risk for any given symptom to last ≥7 days was either lower (chills, headache, rhinorrhea, fatigue) or unchanged (Figure 3; Supplementary Table S4).

**Figure 2.**
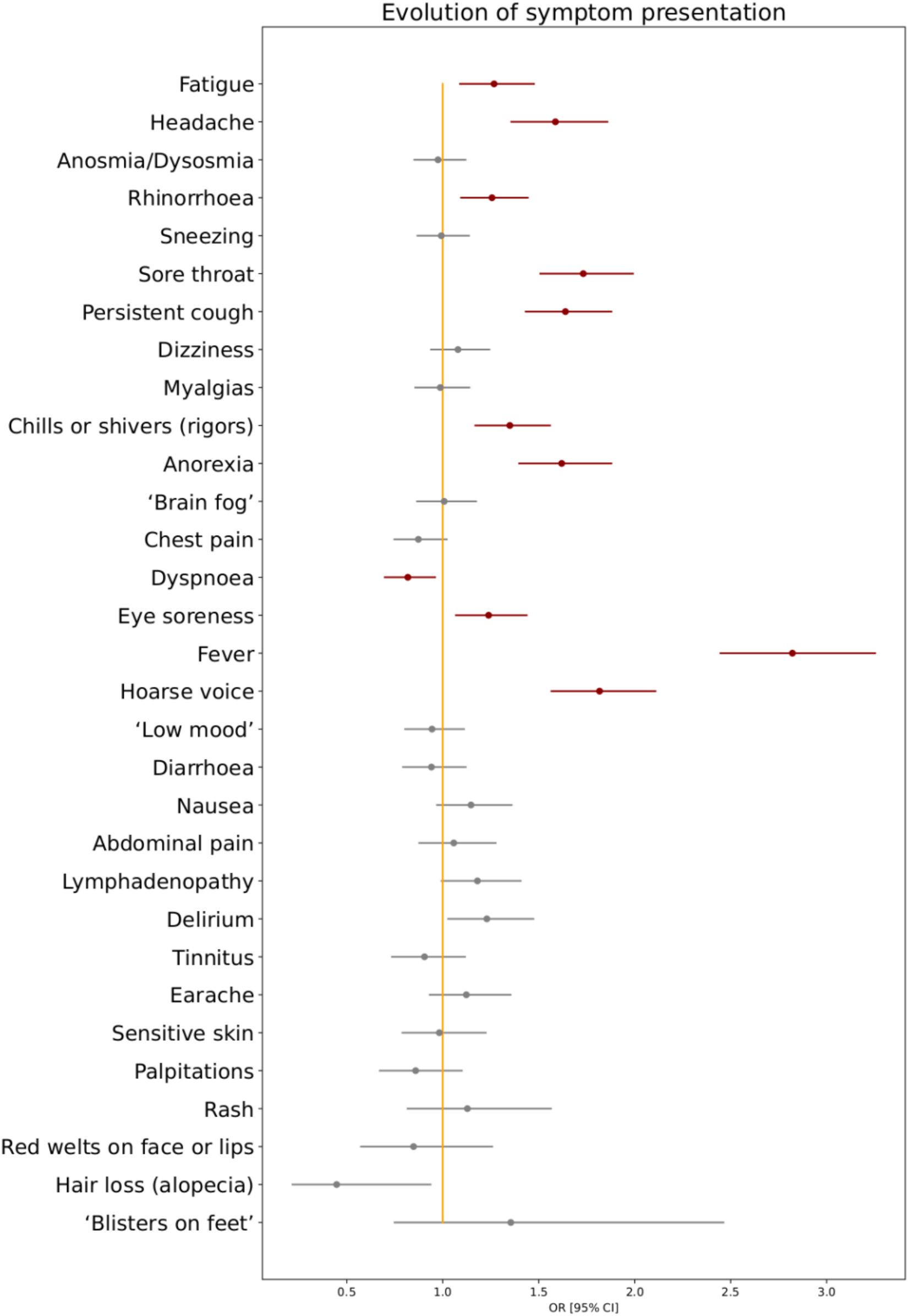
Odds ratios for any symptom presenting within the first 28 days of illness in individuals with COVID-19 during periods of SARS-CoV-2 Delta vs. Alpha variant predominance. Red markers encode statistical significance with α-value < 0·05, whereas grey markers encode non-significant differences, after correction for false discovery rate.

**Figure 3.**
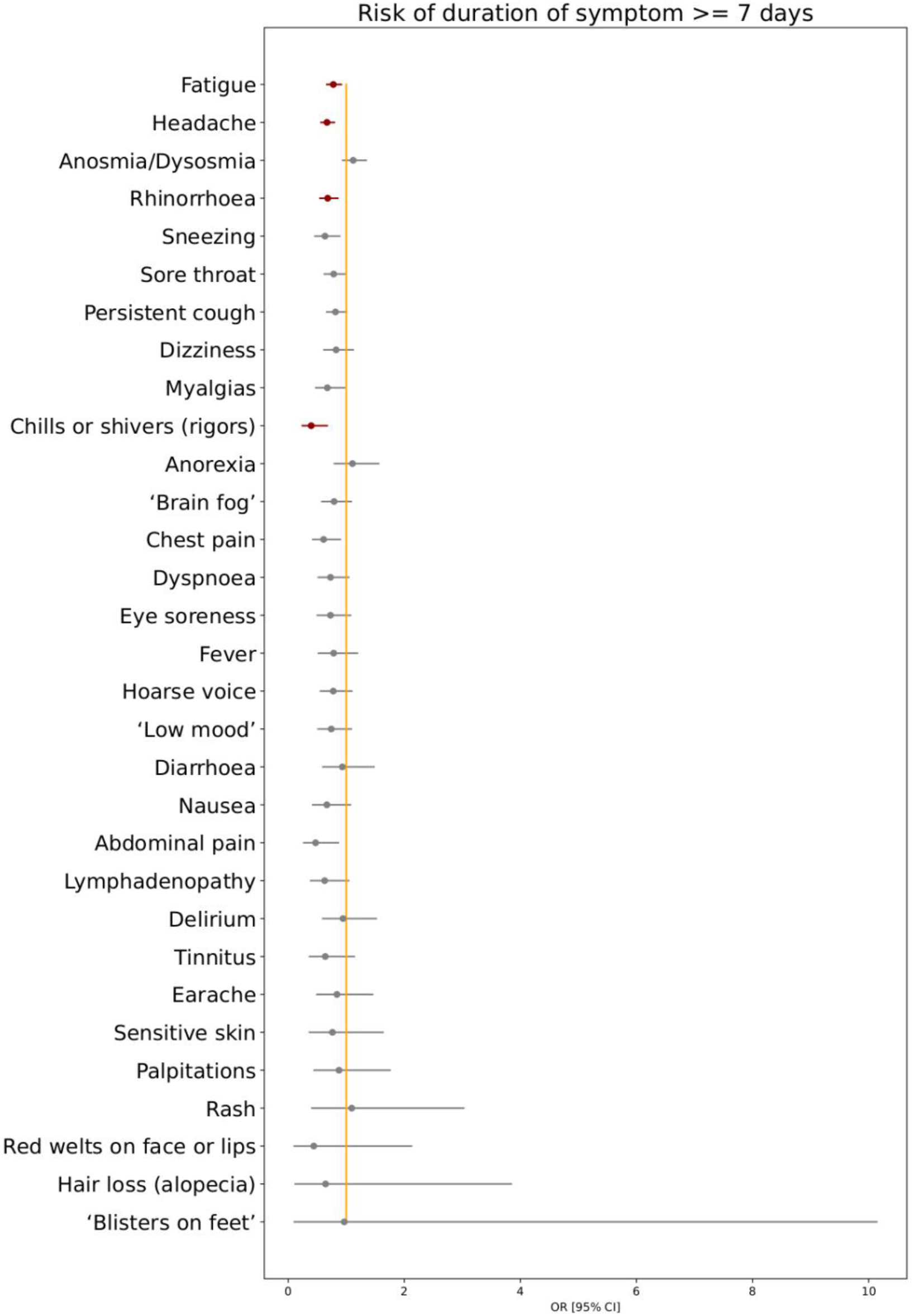
Odds ratios for risk of symptom duration ≥ 7days for individuals with COVID-19 during periods of SARS-CoV-2 Delta vs. Alpha variant predominance. Red markers encode statistical significance with α-value < 0·05 (after FDR correction), whereas grey markers encode non-significant differences.

### Long illness duration

The risk of LC28 was moderately lower with Delta vs. Alpha infection (8·7% [311/5,581] vs. 10·6% [380/5,581] of individuals, OR = 0·69 [0·50; 0·94], p=0·018). Considering only unvaccinated individuals (Table 1), there was a trend towards lowered risk of LC28 with Delta vs. Alpha infection (OR = 0·75 [0·54; 1·04], p= 0·087).

### Hospital presentation

Hospital care for COVID-19 was needed for 120 (3·4%) of 3,581 individuals during the Delta period and 207 (5·8%) of 3,581 during the Alpha period. Noting here that the UK vaccination campaign was stratified by age, clinical vulnerability, and health-care worker status, the risk of hospital presentation was moderately but not significantly lower for Delta vs. Alpha infection, considered overall (OR= 0·76 [95% CI 0·53; 1·11] p=0·156) or for unvaccinated individuals (OR = 0·82 [95% CI 0·56; 1·20], p=0·299).

### Transmissibility

Consistent with other studies, the Delta variant was more transmissible than Alpha, by 1·47 (95%CI 1·45-1·49) according to a population-weighted average (Table 2), noting wide confidence intervals and variation over time. Estimates per region agreed broadly with some regional variation (Figure 4, Table 2).

**Figure 4:**
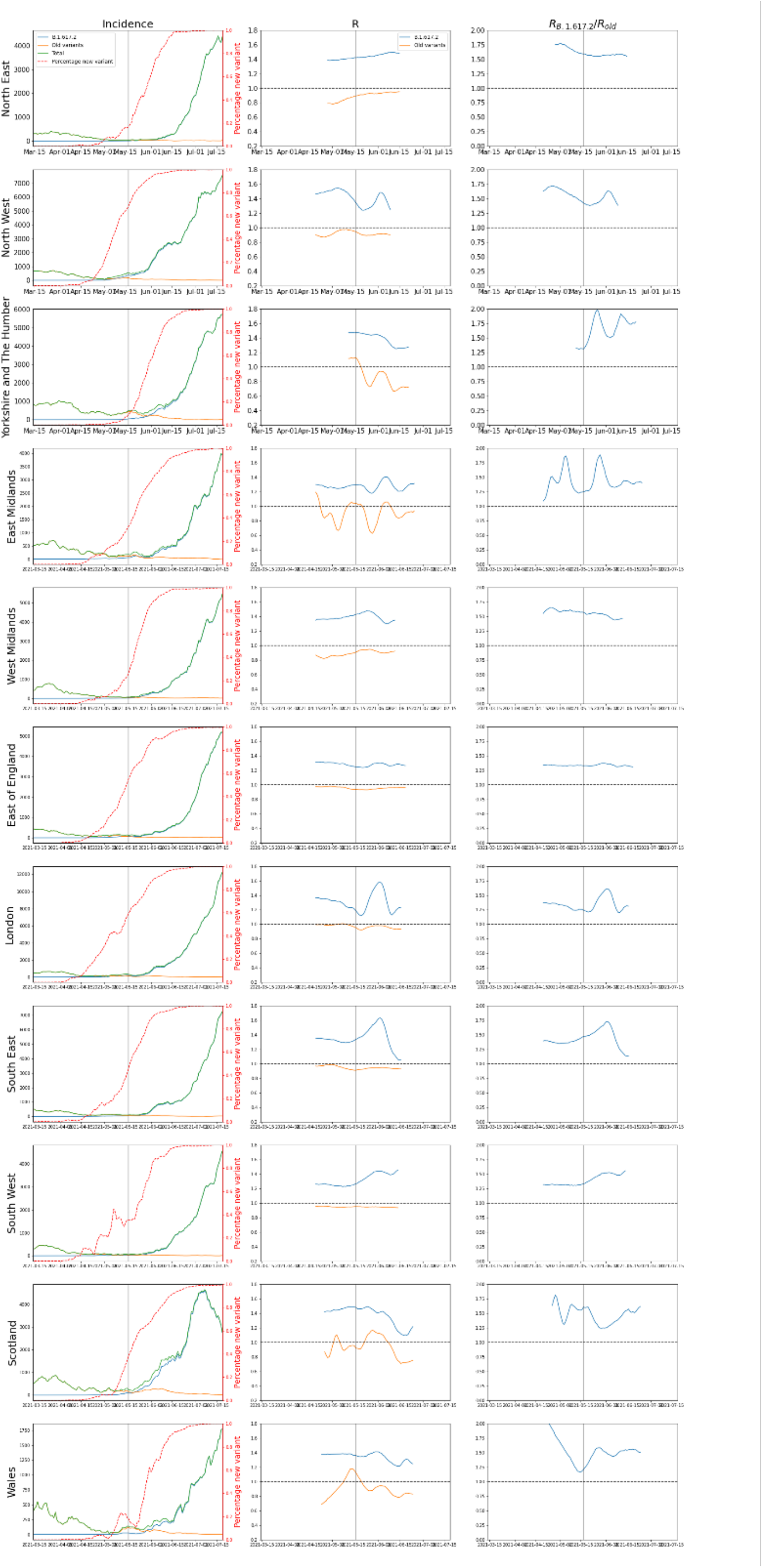
Incidence and R(t) for Delta and non-Delta variants. Left column shows total incidence and incidence for each variant. Middle column shows R(t) for each variant. Rightmost column shows the ratio R_Delta/R_non-Delta, noting that in the timeframe considered non-Delta was predominantly Alpha. Vertical line indicates lifting of some restrictions on May 17, 2021.

**Table 2:** Increase in R(t) for Delta vs. Alpha SARS-CoV-2 variants. R(t) is reported for regions in Great Britain (data from Northern Ireland insufficient to reliably conduct the analysis).

| United Kingdom Region | Multiplicative R increase [mean (95% CI)] |
| --- | --- |
| North East | 1.57 (1.55-1.59) |
| North West | 1.47 (1.31-1.64) |
| Yorkshire and The Humber | 1.69 (1.35-2.03) |
| East Midlands | 1.44 (1.13-1.76) |
| West Midlands | 1.52 (1.43-1.61) |
| East of England | 1.34 (1.30-1.37) |
| London | 1.36 (1.09-1.64) |
| South East | 1.47 (1.11-1.84) |
| South West | 1.47 (1.35-1.59) |
| Scotland | 1.44 (1.18-1.69) |
| Wales | 1.49 (1.31-1.66) |

### Effect of the Delta variant on re-infection

Figure 5 shows the (small) absolute numbers of re-infections across regions, with: a) the number of positive tests reported by app users; and b) the Delta variant as a proportion of circulating SARS-CoV-2, over time. Spearman correlations between reinfection and positive test incidence ranged from 0·46 in the South East to 0·83 in the Midlands. Correlations between reinfection and Delta variant proportions in each region were lower, ranging from 0·41 in the North East and Yorkshire to 0·69 in the North West. In most regions, the correlation of reinfections with the number of reported tests was higher than the correlation of reinfections with the proportion of Delta variant. Supplementary Table S5 presents characteristics of the bootstrapped distribution (100 samples) of correlations for each region over time. Thus, the rise of SARS-CoV-2 infection during the time of Delta predominance correlates more closely with the rise of incidence of new cases *per se*, rather than the rise in proportion of cases due to the Delta variant specifically.

**Figure 5.**
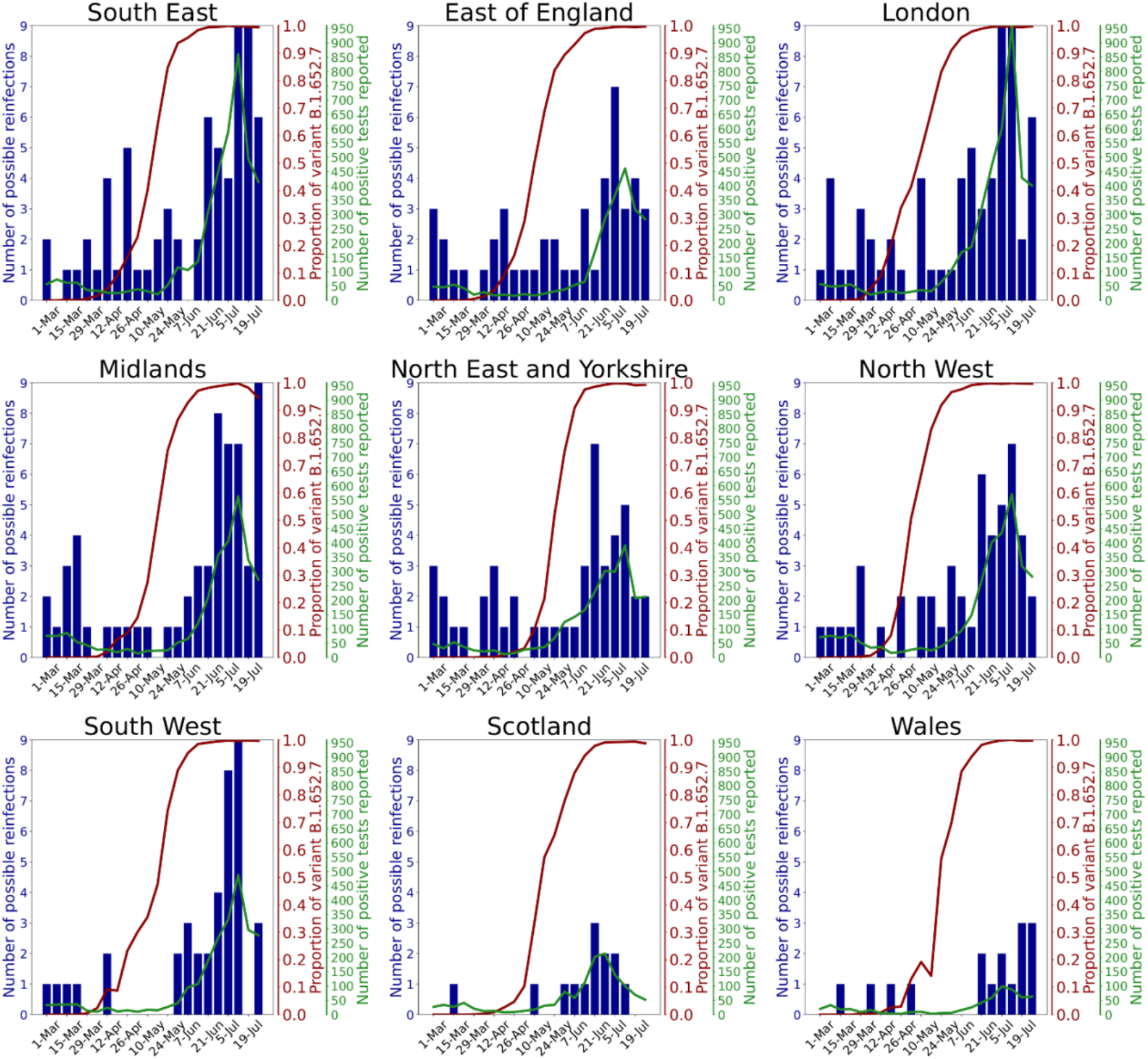
Regional graphs presenting evolution of numbers of reported natural reinfection with SARS-CoV-2. Reported natural reinfection is charted over time in weeks (starting from March 1, 2021) (x-axis). The blue bars graph the absolute number of cases with re-infection. The red line graphs the proportion of the Delta variant among circulating SARS-CoV-2 (COG UK – Supplementary Table S1). The green line graphs the total numbers of positive tests reported by ZOE COVID Symptom Study app users. Data are combined for East and West Midlands, and for Yorkshire and the North East.

### Post-vaccination infection during the Delta period

402,191 app users aged 20-65 years were vaccinated with BNT162b2 (1^st^ dose only: 33,171, 2^nd^ dose: 117,091) or ChAdOx1 nCoV-19 (1^st^ dose only: 59,663, 2^nd^ dose: 192,266) and reported at least one PCR or LFAT test after vaccination between May 26 and July 1, 2021. A positive result was reported by 1,723 of 92,834 (1·86%) who had received one dose, and 1,722 of 309,357 (0·56%) who had received two doses. Data were compared to 25,395 unvaccinated time-matched participants, in whom 1,361 (5·36%) individuals reported a positive test.

After adjustment for population differences in the vaccinated groups using Poisson regressions as described in Methods, the risk reduction of post-vaccination infection after first dose (considered 14-60 days after first dose) was -71·5% (95%CI: -74·4 to -68·3) with BNT162b2 and -58·3% (95%CI: -63·7 to -52·1) with ChAdOx1 nCoV-19, compared with unvaccinated individuals. The risk reduction was even larger in fully vaccinated individuals (considered 14-60 days after second dose): -84·1% [95%CI: -86·9 to -80·6] with BNT162b2, and -69·6% [95%CI: - 72·9 to -65·9] with ChAdOx1 nCoV-19, compared with unvaccinated controls (Figure 6).

**Figure 6.**
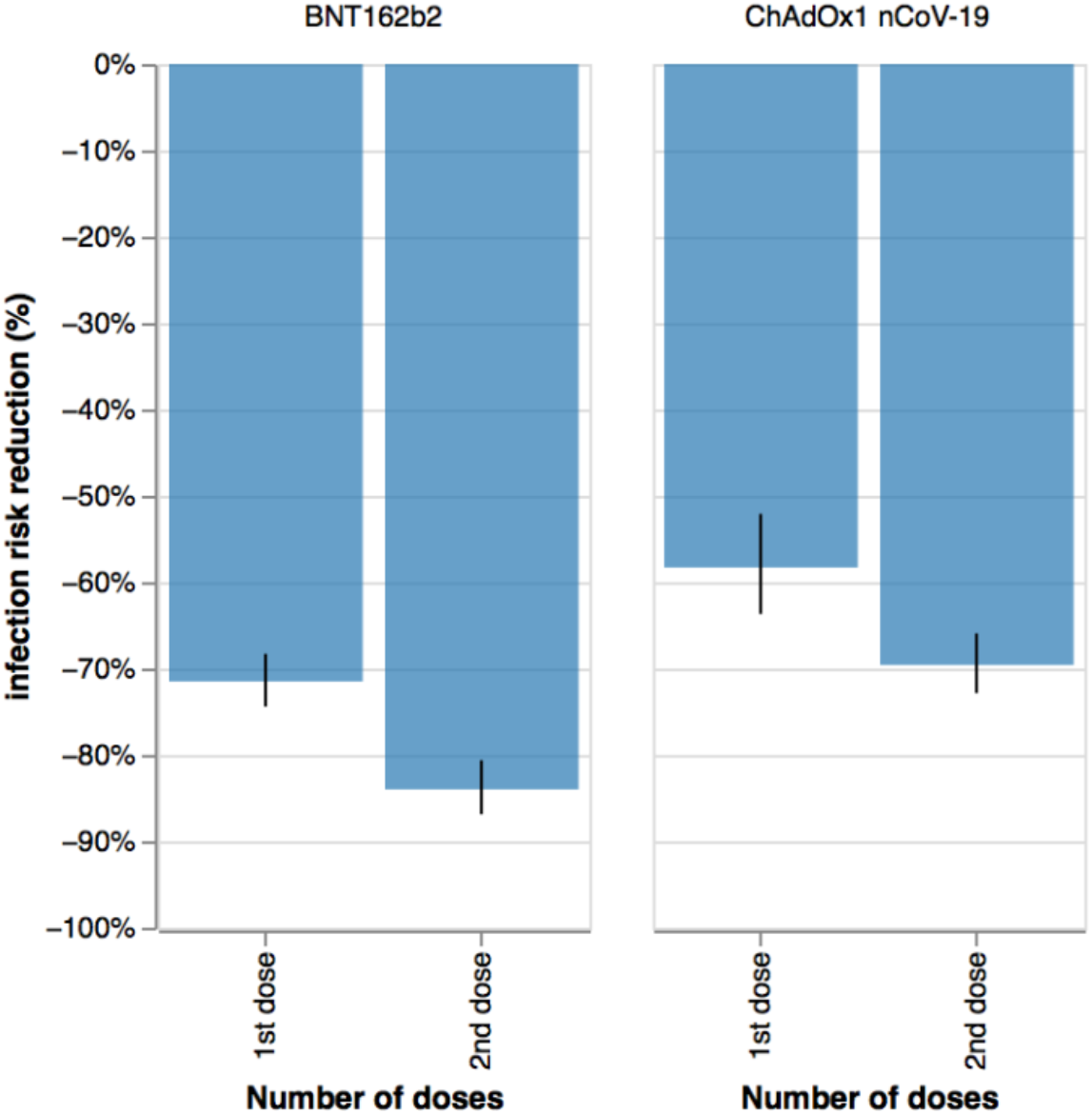
Infection risk reduction (in %) with Delta variant 14-60 days after vaccination after one or two doses of either BNT162b2 or ChAdOx1 nCoV-19 vaccines.

## Discussion

Our large-scale community-based UK study has shown that COVID-19 is clinically similar whether due to Alpha or Delta variants. Ten of 31 symptoms were more common with Delta infection and one with Alpha infection. Although the burden of symptoms in the first week was higher with Delta infection, duration of many individual symptoms was shorter; fewer individuals experienced illness lasting more than 28 days – though saliently this was unchanged in unvaccinated individuals – and there was a trend towards fewer hospital presentations. These observations need to be interpreted in the context of increasing vaccination of the UK population, along with many other environmental and societal changes.

Few studies of COVID-19 due to the Delta variant are available for comparison n. One study of 27 infected young individuals reported symptoms in 22 (81%), with the commonest symptoms fever (41%), cough (33%), headache (26%), and sore throat (26%) (duration of not reported). ^23^ Although there are a few other studies, these all report smaller cohrots. The REACT-1 study Round 14 report (UK data during September 2021, with Delta variant the predominant UK variant) showed a weighted prevalence of individuals testing positive varied greatly by age(0·29% in adults aged >75 years to 2·55% in teenagers and 2·32% in children aged 5-12 years), noting high vaccination rates in older individuals and little or no vaccination in younger age groups at the time of this report.^24^ However, data on symptoms (duration and/or prevalence) were not reported.

The risk of LC28 was lower with Delta (8·7%) vs. Alpha infection (10·6%), although not statistically different in unvaccinated individuals. These results are similar to our previous paper using similar methodology, for individuals infected during the first UK pandemic wave (13·3%).^18^ The Post COVID syndrome (Long COVID) is defined as illness duration >12 weeks after likely SARS-CoV-2 infection (https://www.nice.org.uk/guidance/NG188). Our census dates precludeour ability to compare illness duration beyond 28 days between our two cohorts. Estimates of prevalence of the Post-COVID syndrome are difficult, as many studies lack appropriate control groups. A recent meta-analysis of UK longitudinal cohort studies suggested the post-COVID syndrome was present in 1·2 - 4·8% of individuals,.^25^ similar to recently published figures from the Office for National Statistics (ONS) (1·9% of the UK population self-reporting long COVID (diagnosis otherwise unverified) as of October 2, 2021, although only 71% had (or suspected they had) COVID-19 12 weeks earlier (https://www.ons.gov.uk/peoplepopulationandcommunity/healthandsocialcare/conditionsanddiseases/bulletins/prevalenceofongoingsymptomsfollowingcoronaviruscovid19infectionintheuk/4november2021).

We showed a marked increase in transmissibility with the Delta vs. non-Delta (i.e., Alpha) variant, noting wide confidence intervals. This analysis does not take into account prior natural infection or vaccination rates within the community; and is likely a combination of both the Delta variant’s transmission advantage and its potential ability to evade immunity (whether induced by vaccination or prior natural infection with Alpha or other strains). This estimated increase in transmissibility is greater than we previously estimated for Alpha vs. earlier variants using the same methodology (1·35 (95% CI 1·02–1·69))^15^ noting again pertinent differences (e.g., viral prevalence, lockdown restrictions) between the current and previous studies. Estimates in both studies assume that incidence estimated from app userscan be made representative of the wider population, using stratification by age and vaccination status. However, other factors such as behavior and socio-economic status are not corrected for by this analysis. Other studies have also identified higher Delta transmissibility, resulting in rising incidence particularly in young unvaccinated age groups, higher re-infection rates, and a higher viral load in infected individuals.^26^ Here we note the REACT-1 study report of an exponential increase in infections in children aged 5-17 years in September 2021, coinciding with return-to-school, with most school-age children unvaccinated at this time.

Our study found that for most regions of the UK, the correlation of reinfections with the number of reported positive tests (i.e., incidence of cases) was higher than the correlation of reinfections with the proportion of Delta among the circulating variants (i.e., incidence of variant). In other words, the rise in COVID-19 correlated more closely with increase in prevalence of SARS-CoV-2 overall rather than the increased proportion of circulating SARS-CoV-2 due to the Delta variant. SARS-CoV-2 infection provides substantial and persistent immunologic protection for at least several months for most individuals, with a recent systematic review suggesting a risk reduction of reinfection of >90%, similar to vaccination, and evident for at least 10 months.^32^. However, this may not be uniform across the population. A study of tested individuals followed prospectively for at least 3 months demonstrated a protective effect after prior infection of 80·3% for younger individuals (aged 20-59 years) but only 67·4% for older individuals (aged ≥60 years), with lower levels of protection in individuals associated with a long-term care facility and/or who had milder initial disease.^27^ A study of Danish healthcare workers^28^ found small absolute numbers of re-infected individuals (5 of 750 seropositive individuals over 5-6 months). However, 5% of previously seropositive individuals reverted to seronegative status, associated with older age and fewer symptoms with initial infection (noting here that the relationship between antibody titre and subsequent infection risk is currently unclear). In July 2021 UK governmental figures estimated an adjusted odds ratio of reinfection risk from the Delta variant vs. the Alpha variant as 1·5. However, this varied according to time since initial infection: the odds ratio was not elevated if initial infection was <180 days earlier (adjusted odds ratio = 0·8), but was higher if initial infection was ≥180 days earlier (adjusted odds ratio = 2·4) (https://assets.publishing.service.gov.uk/government/uploads/system/uploads/attachment_data/file/1005517/Technical_Briefing_19.pdf).

Our observational data support effectiveness of both BNT162b2 and ChAdOx1 nCoV-19 vaccines against the Delta variant. Both reduced the risk of testing positive during the Delta period, evident after the first and enhanced after the second dose.^29^ These figures are similar to our previous results when the Alpha variant was predominant.^15^ We have an inherent bias due to the nature of the UK vaccine rollout, whereby health-care workers, elderly people, and clinically vulnerable individuals were prioritised before the younger population, creating unbalanced demographic characteristics between vaccinated and unvaccinated populations. Moreover, we cannot compare vaccination effectiveness against Alpha vs. Delta variants, given the many differences between the two timeframes. Although we attempted to adjust for some of these differences using Poisson regression, behavioural factors are difficult to capture. For example, individuals vaccinated earlier may have changed their behaviours over concern of possible waning antibody status and possible reducing immunity.^30^ However, our results concord with vaccination trial data,^31,32^ and provide support for ongoing vaccination campaigns internationally. Previous data have shown that vaccination is associated with significant reduction in risk of hospitalisation and disease progression to death or mechanical ventilation in individuals with COVID-19.^12,33^ Our data similarly showed a trend towards fewer hospital presentations. A later analysis during the UK’s third wave will be useful here.

We acknowledge the limitations of our observational study. Self-reported data from a mobile phone app may disproportionately represent more affluent populations and can introduce information bias and/or effect bias, although previous work from the CSS has shown that our self-reported data aligns well with surveys designed to be representative of the population.^34^ Participants could only report a positive test and we cannot confirm the actual variant causing infection, although our assumptions of Delta and Alpha infection are strongly supported by UK-COG surveillance variant testing. During the study, both overall numbers and individual app users fluctuated in their participation in the CSS-app, potentially for many factors including mass-media information, summer vacation, and perception of relevance. Our populations were matched for age and sex but not BMI; and we note higher diabetes prevalence and BMI in the Alpha cohort. Relevantly, vaccination was not only tiered by age but also to those with co-morbidities including diabetes. As mentioned, the timeframes of Alpha and Delta variant predominance differed with respect to guidance on social distancing and behavior in public spaces, highly likely to affect viral diffusion in the population, thus affecting our transmission calculations. Last, vaccine effectiveness could only be determined in tested individuals, noting that we do not have information regarding the reason for testing in these individuals. We were also only able to assess individuals in the age range of 20-65, in order to avoid unbalanced case/control data. Relevantly, early post-vaccination symptoms can mimic COVID-19^35^ but may not necessarily trigger testing. Here, our previous work showed that vaccinated individuals are more likely to have post-vaccination systemic symptoms after a previously positive testcompared to those without known past infection (odds ratios 2·3 - 4·0), which may bias presentation for SARS-CoV-2 testing post-vaccination.

## Conclusions

The clinical presentation of COVID-19 due to the Delta variant is similar to illness caused by the Alpha variant: although symptom burden in the first week is modestly higher, individual symptom duration was either the same or shorter, and the risk of LC28 was lower. The Delta variant was more transmissible than the preceding predominant variant (i.e., Alpha) but did not increase the risk of reinfection *per se*. The risk of infection in fully vaccinated individuals was reduced by both BNT162b2 and ChAdOx1 nCoV-19, compared with unvaccinated controls, confirming good vaccine efficacy against the Delta variant and supporting prosecution of the anti-SARS-CoV-2 vaccination campaign internationally.

## Data Availability

Data collected in the COVID Symptom Study smartphone app are being shared with other health researchers through the UK NHS-funded Health Data Research UK and Secure Anonymised Information Linkage consortium, housed in the UK Secure Research Platform (Swansea, UK). Anonymised data are available to be shared with researchers according to their protocols in the public interest (https://web.www.healthdatagateway.org/dataset/fddcb382-3051-4394-8436-b92295f14259).

## Abbreviations

CI: confidence interval
COVID-19: coronavirus disease 2019
IQR: inter-quartile range
LC28: long COVID-19 with duration of symptoms ≥28 days
LFAT: lateral flow antigen test
ONS: Office for National Statistics (UK)
PCR: polymerase chain reaction
SARS-CoV-2: severe acute respiratory syndrome-related coronavirus 2
UK: United Kingdom of Great Britain and Northern Ireland
WHO: World Health Organisation

## ACKNOWLEDGEMENTS

This research was funded in part by the Wellcome Trust (WT213038/Z/18/Z). This work is also supported by the Wellcome Engineering and Physical Sciences Research Council Centre for Medical Engineering at King’s College London (WT203148/Z/16/Z) and the UK Department of Health via the National Institute for Health Research (NIHR) comprehensive Biomedical Research Centre award to Guy’s and St Thomas’ NHS Foundation Trust in partnership with King’s College London and King’s College Hospital NHS Foundation Trust, the Medical Research Council (MRC), and British Heart Foundation. SO and MM are supported by the UK Research and Innovation London Medical Imaging and Artificial Intelligence Centre for Value Based Healthcare, and the Wellcome Flagship Programme (WT213038/Z/18/Z). EM is funded by an MRC Skills Development Fellowship Scheme at King’s College London. CHS is supported by the National Core Studies, an initiative funded by United Kingdom Research and Innovation, NIHR, and the Health and Safety Executive, and funded by MRC (MC_PC_20030). CHS is also supported by an Alzheimer’s Society Junior Fellowship (ASJF-170–11). ZOE Limited supported all aspects of building and running the application and service to all users worldwide. For the purpose of open access, the authors have applied a CC BY public copyright licence to any Author Accepted Manuscript version arising from this submission. COG-UK is supported by funding from the Medical Research Council (MRC) part of UK Research & Innovation (UKRI), the National Institute of Health Research (NIHR) [grant code: MC_PC_19027], and Genome Research Limited, operating as the Wellcome Sanger Institute. The authors are grateful to Michael Absoud and Sunil Bhopal for constructive comments on the paper.

