## Supplemental Material for "COVID-19 due to the B.1.617.2 (Delta) variant compared to B.1.1.7 (Alpha) variant of SARS-CoV-2: two prospective observational cohort studies"

### Supplementary Methods

The COVID Symptom Study (administered through the ZOE COVID Study App) was launched jointly by ZOE Limited and King's College London on March 24, 2020. Data were acquired through a mobile application.[17] Participants can log data for themselves and can proxy-report for other individuals (e.g., spouses, elderly relatives).

At time of registration through the app, all participants provide consent for their data to be used for research. Participants can withdraw from the study at any time, with their data, (including proxy-reported data) subsequently excluded from analysis.

The app was tested both before launch and before version updates. The app usage workflow was described in the earliest publication about the COVID Symptom Study (doi: 10.1126/science.abc0473). The team of app developers monitor the statistics of app usage and users' input daily, to assess for faults (e.g., problems in the app downloads or logging). No major disruptions were detected during this study. Although the questions asked through the app have changed over time, the last change to COVID-19 symptom direct questioning was 4 November 2020 (Supplementary Table S2); thus all symptom questions were asked of all individuals during both time periods reported in the current study.

Conjointly to the app development, software for data extraction, curation and analytics was developed, which guarantees the repeatability of the data curation over time and the consistency on different timestamps, according to stringent predefined criteria (filters). The software, based on Python language, is called ExeTera;[36] documentation and associated code can be downloaded here: https://arxiv.org/abs/2011.00867

Currently 92% of UK adults own a smartphone, with little difference according to socioeconomic status (https://www.statista.com/statistics/300384/mobile-phoneusage-in-the-uk-by-socio-economic-group/).

**Supplementary Table S1. Incidence (%) of Alpha and Delta variants of SARS-CoV-2 in United Kingdom, in the weeks starting 30 December 2020 - 5 May 2021 and 26 May - 7 July 2021**

(https://assets.publishing.service.gov.uk/government/uploads/system/uploads/attachment_data/file/975754/Variants_of_Concern_Technical_Briefing_8_Data_England.xlsx; GISAID https://covariants.org).

| Week starting date | Alpha (B.1.1.7) | Delta (B.1.617.2) |
|---|---|---|
| 30 December 2020 | 78·1 | 0 |
| 6 January | 82·3 | 0 |
| 13 January | 87·6 | 0 |
| 20 January | 90·8 | 0 |
| 27 January | 93·8 | 0 |
| 3 February | 95·7 | 0 |
| 10 February | 97·4 | 0 |
| 17 February | 98·1 | 0 |
| 24 February | 98·6 | 0 |
| 3 March | 98·8 | 0 |
| 10 March | 98·9 | 0 |
| 17 March | >99 | 0 |
| 24 March | 98 | 0 |
| 31 March | 98 | 0 |
| 7 April | 95 | 2 |
| 14 April | 92 | 6 |
| 21 April | 87 | 9 |
| 28 April | 76 | 24 |
| 5 May | 71·5 | 25·8 |
| 26 May | 19·3 | 80·0 |
| 2 June | 9·4 | 90·2 |
| 9 June | 5 | 95 |

| | | |
|---|---|---|
| 16 June | 2 | 98 |
| 23 June | 2 | 98 |
| 30 June | <1 | >99 |
| 7 July | <1 | >99 |

**Supplementary Table S2. List of symptom questions asked by the ZOE Symptom Study app after 4 November 2020.**

| **Symptom** | **COVID Symptom Study app question** |
|---|---|
| Fever | Fever (at least 37·8C or 100F) |
| Persistent Cough | Persistent cough (coughing a lot for more than an hour or 3 or more coughing episodes in 24 hours) |
| Fatigue | Unusual fatigue… (no; mild fatigue; severe fatigue/ I struggle to get out of bed) |
| Dyspnoea | Shortness of breath or trouble breathing (no; yes mild symptoms/ slight shortness of breath during ordinary activity: yes significant symptoms/ breathing is comfortable only at rest; yes, severe symptoms/ breathing is difficult even at rest). |
| Anosmia/Dysosmia | Loss of smell |
| Ageusia | Loss of taste |
| Hoarse Voice | Unusually hoarse voice |
| Chest Pain | Unusual chest pain or tightness in your chest |
| Abdominal Pain | Unusual abdominal pain or stomach-ache |
| Diarrhoea | Diarrhoea |
| Stools | How many loose stools in the last 24 hours? |
| Headache Frequency | How often are you experiencing headaches? |
| Delirium | Confusion, disorientation or drowsiness |
| Eye Soreness | Do your eyes have any unusual eye-soreness or discomfort (e.g. light sensitivity, excessive tears, or pink/red eye)? |
| Anorexia | Skipping meals |
| Headache | Headache |
| Nausea | Nausea or vomiting |
| Dizziness | Dizziness or light-headedness |

| | |
|---|---|
| Sore Throat | Sore or painful throat |
| Myalgias | Unusual strong muscle pains or aches |
| Red Welts | Raised, red, itchy welts on the skin or sudden swelling of the face or lips |
| Blisters | Red/purple sores or blisters on your feet, including your toes |
| Allergy Exacerbation | Increase in your usual allergy symptoms |
| Rashes | Rash on your arms or torso |
| ‘Sensitive Skin’ | Strange, unpleasant sensations in your skin like pins & needles or burning |
| Hair Loss (alopecia) | Unusual hair loss |
| Low Mood | Feeling down, depressed or hopeless |
| Brain Fog | Loss of concentration or memory (brain fog) |
| Dysosmia | Altered smell (things smell different to usual) |
| Dysgeusia | Altered taste (things taste different to usual) |
| Rhinorrhoea | Runny nose |
| Sneezing | Sneezing more than usual |
| Ear Pain | Earache |
| Tinnitus | Ringing in your ears |
| Lymphadenopathy | Swollen neck glands |
| Palpitations | Unusually fast or irregular heartbeat (palpitations) |
| Arthralgias | Unusual joint pains or aches |
| Mouth Ulcers | Mouth or tongue ulcers |
| Tongue Changes | Changes to tongue surface |

**Supplementary Table S3. Number of individuals presenting with a given symptom over the course of the disease, ranked according to the order during the Alpha period.** Symptom onset between December 28, 2020 and May 6, 2021 was attributed to infection with the Alpha variant, and symptom onset between May 26, 2021 and July 1, 2021, to infection with the Delta variant.

| | **Alpha**<br>(n=3,581)<br>N (%) | **Delta**<br>(n= 3,581)<br>N (%) |
|---|---|---|
| **Fatigue** | 2,613 (73·0) | 2625 (73·3) |
| **Headache** | 2,387 (66·7) | 2,668 (74·5) |
| **Anosmia/Dysosmia** | 2,190 (61·2) | 2,287 (63·9) |
| **Rhinorrhoea** | 1,935 (54·0) | 2,557 (71·4) |
| **Sneezing** | 1,564 (43·7) | 2,111 (59·0) |
| **Sore throat** | 1,515 (42·3) | 2,022 (56·5) |
| **Persistent cough** | 1,457 (40·7) | 1,813 (50·6) |
| **Dizziness** | 1,233 (34·4) | 1,281 (35·8) |
| **Myalgias** | 1,135 (31·7) | 1,010 (28·2) |
| **Chills or shivers (rigors)** | 1,026 (28·7) | 1,260 (35·2) |
| **Anorexia** | 996 (27·8) | 1,015 (28·3) |
| **'Brain fog'** | 922 (25·8) | 1,064 (29·7) |
| **Chest pain** | 912 (25·5) | 778 (21·7) |
| **Dyspnoea** | 908 (25·4) | 774 (21·6) |
| **Eye soreness** | 901 (25·2) | 1041 (29·1) |
| **Fever** | 883 (24·7) | 1548 (43·2) |
| **Hoarse voice** | 844 (23·6) | 1286 (35·9) |
| **'Low mood'** | 843 (23·5) | 709 (19·8) |

| | | |
|---|---|---|
| **Diarrhoea** | 759 (21·2) | 663 (18·5) |
| **Nausea** | 719 (20·1) | 641 (17·9) |
| **Abdominal pain** | 570 (15·9) | 528 (14·7) |
| **Lymphadenopathy** | 565 (15·8) | 701 (19·6) |
| **Tinnitus** | 502 (14·0) | 559 (15·6) |
| **Delirium** | 502 (14·0) | 543 (15·2) |
| **Earache** | 482 (13·5) | 626 (17·5) |
| **Sensitive skin** | 427 (11·9) | 334 (9·3) |
| **Palpitations** | 324 (9·1) | 318 (8·9) |
| **Rash** | 167 (4·7) | 154 (4·3) |
| **Red welts on face or lips** | 132 (3·7) | 90 (2·5) |
| **Hair loss (alopecia)** | 44 (1·2) | 43 (1·2) |
| **'Blisters on feet'** | 36 (1·0) | 46 (1·3) |

**Supplementary Table S4. Risk of a given symptom over the first 28 days of illness, and of a symptom lasting for at least 7 days, comparing Delta with Alpha infection.** Bold font indicates that the significance survives FDR correction.

| | Risk of symptom within first 28 days of illness | | When present, risk of symptom lasting at least 7 days (note: all symptoms presented by Day 21 of illness) | |
|---|---|---|---|---|
| | **OR (95%CI)** | **p-value** | OR (95%CI) | **p-value** |
| **Fatigue** | **1·27 [1·09 - 1·48]** | **0·003** | **0·78 [0·65 - 0·93]** | **0·005** |
| **Headache** | **1·59 [1·35 - 1·86]** | **<0·001** | **0·67 [0·55 - 0·81]** | **<0·001** |
| Anosmia / Dysosmia | 0·98 [0·85 - 1·12] | 0·728 | 1·12 [0·92 - 1·36] | 0·246 |
| **Rhinorrhoea** | **1·26 [1·09 - 1·45]** | **0·002** | **0·68 [0·53 - 0·87]** | **0·002** |
| **Sore throat** | **1·73 [1·50 - 2·00]** | **<0·001** | 0·78 [0·61 - 1·01] | 0·056 |
| Sneezing | 0·99 [0·86 - 1·14] | 0·915 | 0·63 [0·45 - 0·90] | 0·011 |
| **Persistent cough** | **1·64 [1·43 - 1·88]** | **<0·001** | 0·81 [0·65 - 1·02] | 0·075 |
| Dizziness | 1·08 [0·93 - 1·25] | 0·302 | 0·82 [0·60 - 1·13] | 0·233 |
| **Chills or shivers (rigors)** | **1·35 [1·17 - 1·56]** | **<0·001** | **0·40 [0·23 - 0·69]** | **0·001** |
| Myalgias | 0·99 [0·85 - 1·14] | 0·864 | 0·67 [0·46 - 0·99] | 0·042 |
| **Fever** | **2·82 [2·44 - 3·26]** | **<0·001** | 0·78 [0·51 - 1·21] | 0·266 |
| **Anorexia** | **1·62 [1·39 - 1·88]** | **<0·001** | 1·11 [0·78 - 1·57] | 0·566 |
| 'Brain fog' | 1·01 [0·86 - 1·18] | 0·927 | 0·79 [0·57 - 1·10] | 0·162 |
| **Eye soreness** | **1·24 [1·06 - 1·44]** | **0·006** | 0·73 [0·49 - 1·09] | 0·121 |
| **Hoarse voice** | **1·82 [1·56 - 2·11]** | **<0·001** | 0·78 [0·54 - 1·11] | 0·166 |
| Chest pain | 0·87 [0·74 - 1·03] | 0·099 | 0·61 [0·41 - 0·91] | 0·016 |
| **Dyspnoea** | **0·82 [0·69 - 0·96]** | **0·017** | 0·73 [0·50 - 1·06] | 0·094 |
| 'Low mood' | 0·94 [0·80 - 1·12] | 0·497 | 0·74 [0·50 - 1·10] | 0·138 |
| Diarrhoea | 0·94 [0·79 - 1·12] | 0·502 | 0·93 [0·58 - 1·49] | 0·772 |
| Nausea | 1·15 [0·97 - 1·36] | 0·118 | 0·67 [0·41 - 1·09] | 0·102 |
| Abdominal pain | 1·06 [0·87 - 1·28] | 0·568 | 0·47 [0·25 - 0·88] | 0·018 |
| Lymphadenopathy | 1·18 [0·99 - 1·41] | 0·067 | 0·63 [0·37 - 1·06] | 0·079 |
| Delirium | 1·23 [1·02 - 1·48] | 0·027 | 0·94 [0·58 - 1·53] | 0·815 |
| Earache | 1·12 [0·93 - 1·36] | 0·233 | 0·84 [0·48 - 1·47] | 0·539 |
| Tinnitus | 0·90 [0·73 - 1·12] | 0·359 | 0·64 [0·35 - 1·15] | 0·136 |

| | | | | |
|---|---|---|---|---|
| Sensitive skin | 0·98 [0·79 - 1·23] | 0·876 | 0·76 [0·35 - 1·65] | 0·490 |
| Palpitations | 0·86 [0·67 - 1·10] | 0·236 | 0·88 [0·43 - 1·77] | 0·712 |
| Rash | 1·13 [0·81 - 1·57] | 0·472 | 1·09 [0·39 - 3·04] | 0·863 |
| Red welts on face or lips | 0·85 [0·57 - 1·26] | 0·416 | 0·44 [0·09 - 2·14] | 0·309 |
| Hair loss | 0·45 [0·21 - 0·94] | 0·034 | 0·64 [0·11 - 3·86] | 0·630 |
| 'Blisters' on feet | 1·35 [0·74 - 2·47] | 0·321 | 0·97 [0·09 - 10·16] | 0·977 |

**Supplementary Figure 1. Presence of Delta variant, as a proportion of total SARS-COV-2 positive tests sequenced at that time within England (nine regions), Scotland, Wales and Northern Ireland, using genomic surveillance data (COG-UK).** Grey bars indicate the number of samples sequenced by COG-UK daily. The solid red line shows the proportion of sequenced samples that were identified as Delta variant vs. the total sequenced samples, over time.

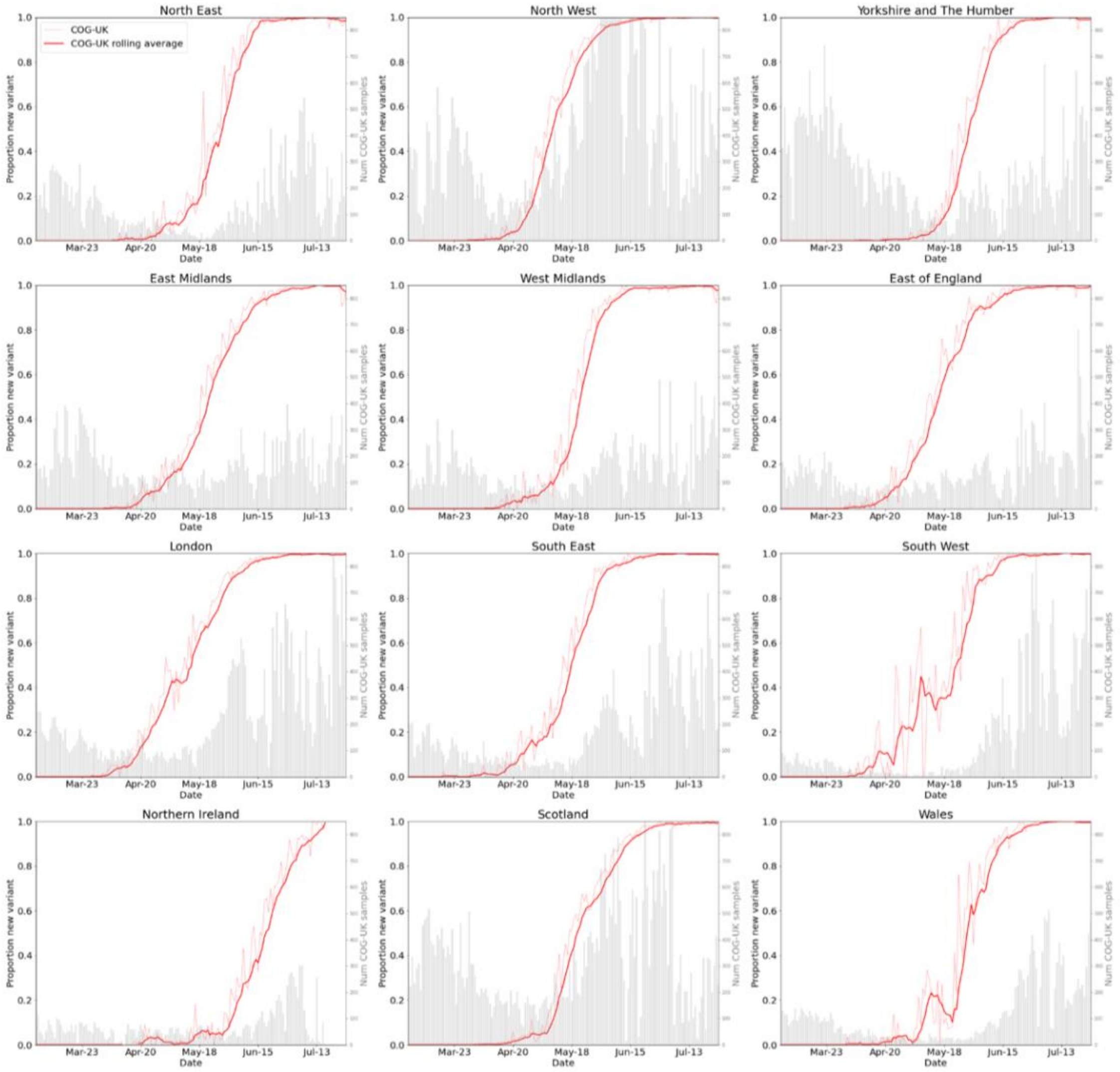

**Supplementary Table S5. Spearman correlations of incidence and variant proportion with reinfection (both direct and bootstrapped data).**

Comparison of these two distributions of correlations in each region, using the Mann-Whitney U test (using bootstrapped data).

| | **Spearman correlation of number of positive tests and numbers of reinfections** | | **Spearman correlation of proportion of Delta variant [as proportion of total circulating SARS-CoV-2) and numbers of reinfections** | | |
|---|---|---|---|---|---|
| **UK Region** | **Direct** | **Bootstrapped (median [IQR])** | **Direct** | **Bootstrapped (median [IQR])** | **Comparison p-value** |
| South East | 45·83 | 45·85 [31·55; 58·47] | 65·14 | 62·08 [51·57; 72·28] | <0·0001 |
| East of England | 58·20 | 58·90 [48·97; 69·78] | 50·77 | 52·86 [40·90; 62·00] | 0·002 |
| London | 62·24 | 63·07 [51·42; 72·86] | 57·89 | 59·46 [50·38; 70·17] | 0·099 |
| Midlands | 85·43 | 82·76 [78·57; 86·28] | 49·93 | 44·41 [32·90; 57·31] | <0·0001 |
| North East and Yorkshire | 52·85 | 50·68 [38·72; 61·97] | 42·85 | 41·04 [28·35; 53·32] | <0·0001 |
| North West | 61·81 | 64·65 [55·93; 72·67] | 68·48 | 68·71 [60·93; 75·45] | 0·008 |
| South West | 79·05 | 80·30 [71·48; 88·94] | 47·16 | 44·34 [30·72; 59·87] | <0·0001 |
| Scotland | 68·95 | 65·77 [53·25; 75·66] | 51·26 | 47·52 [33·25; 60·77] | <0·0001 |
| Wales | 59·39 | 63·09 [54·29; 69·18] | 51·66 | 53·41 [37·61; 64·98] | <0·0001 |
